# The 1968 Influenza Pandemic and COVID-19 Outcomes

**DOI:** 10.1101/2021.10.23.21265403

**Authors:** Charles A Taylor, Christopher Boulos, Matthew J. Memoli

## Abstract

Past pandemic experience can affect health outcomes in future pandemics. This paper focuses on the last major influenza pandemic in 1968 (H3N2), which killed up to 100,000 people in the US. We find that places with high influenza mortality in 1968 experienced 1-4% lower COVID-19 death rates. Our identification strategy isolates variation in COVID-19 rates across people born before and after 1968. In places with high 1968 influenza incidence, older cohorts experience lower COVID-19 death rates *relative* to younger ones. The relationship holds using county and patient-level data, as well as in hospital and nursing home settings. Results do not appear to be driven by systemic or policy-related factors, instead suggesting an individual-level response to prior influenza pandemic exposure. The findings merit investigation into potential biological and immunological mechanisms that account for these differences—and their implications for future pandemic preparedness.

## Introduction

The COVID-19 pandemic resulted in the global implementation of an unprecedented set of public health and economic measures including social distancing and supply chain innovations, along with widespread emergency use of new pharmaceuticals and vaccines. To evaluate pandemic response effectiveness and to better understand how to prepare for and prevent future pandemics, researchers are investigating the epidemiological and pathological nature of the SARS-CoV-2 virus in the hope of fully assessing the disease’s global impact—and, in particular, investigating the susceptibility of populations to illness. This paper presents novel findings associating distant life experiences—and potentially biological factors—with the severity of COVID-19 illness, and contributes to several strands of related research spanning the social and biological sciences.

One such research area involves the investigation of heterogeneous COVID-19 outcomes among different populations. Previous work has presented evidence linking demographic, racial, social, and policy factors to the spread and severity of COVID-19 (Dowd et al. 2020; Kraemer et al. 2020; Wells et al. 2020; Taylor et al. 2020; Knittel and Ozaltun 2020; Nepomuceno et al. 2020; Papageorge et al. 2021). Pandemic responses have also varied in terms of mitigation behavior, both globally and within the US, and even under similar governmental policies and regulations. It has been proposed that such differential responses can explain to varying COVID-19 outcomes, but these relationships have been difficult to demonstrate clearly. Our paper contributes to this strand of empirical research, which sits largely within the social sciences, by proposing an additional immunological factor that may confound results of existing studies.

A second relevant area of research explores genetic, immunological, and other biological factors linked to the severity and transmissibility of COVID-19 (Lauc and Sinclair 2020; Hou et al. 2020; Sette and Crotty 2020; Doshi 2020). Such research includes genomic association studies, an investigation of sex differences in immunity, and a proposal that a major genetic risk factor can be traced to Neanderthal DNA (Group 2020; Takahashi and Iwasaki 2021; Zeberg and Pääbo 2020). This category of work explores whether risk factors, biomarkers, and cross-reactivities can be utilized to develop best practices in prophylaxis, testing, and management. Our study may motivate further work in this area by presenting empirical population-level findings potentially pointing to a biological source of COVID-19 resistance.

A third related research thread explores the persistence of past public crises and how they alter human and institutional responses in such a way that mediates the spread, morbidity, and mortality of COVID-19 (Huang et al. 2020; Lokshin et al. 2020). These studies propose previously unexplored mechanisms that contribute to illness, rather than focusing on a specific factor. For instance, Lokshin et al. found higher WWII mortality to be associated with lower COVID-19 death rates, positing a link between institutional investment, social capital, and pandemic response. Other studies have linked SARS exposure to lower COVID-19 mortality, which some have attributed to mask-wearing norms that made compliance with public health measures ‘easier’ (Peter Zhixian Lin and Meissner 2020; Cheng et al. 2020; Lo and Hsieh 2020). Relatedly, our study explores how societal responses to a past public health crisis may explain phenomena occurring several generations later.

Looking beyond COVID-19, there is abundant research into the links between pandemics and disease. One representative study connects an HIV-resistant gene to exposure to the plague and smallpox (Galvani and Slatkin 2003; Sabeti et al. 2005). Another study proposes that the 1918 influenza pandemic hastened the decline of tuberculosis (Noymer and Garenne 2000). Others have linked excess youth mortality during the 1918 influenza pandemic to exposure to the 1889 ‘Russian Flu’ virus (Worobey et al. 2014; Abildgren 2021).

Our paper applies a similar lens to an underexplored but arguably consequential pandemic, the 1968 influenza pandemic, which killed up to 100,000 people in the US. We find that areas with high influenza mortality in 1968 experienced 1-4% lower COVID-19 death rates. Leveraging variation in COVID-19 rates across people born before and after 1968, we find that in areas with high 1968 influenza incidence, older cohorts experience lower COVID-19 death rates *relative* to younger ones. The findings are robust to using county-and patient-level data, as well as hospital and nursing home data, and do not appear to be driven by systemic or policy-related factors. Instead, they suggest a potential biological and immunological response to prior influenza pandemic exposure.

### The 1968 influenza pandemic

While many have looked to the 1918 influenza pandemic for insight into the ongoing epidemiological and economic effects of the COVID-19 pandemic (Beach et al. 2020; Peter Z Lin and Meissner 2021), fewer have explored a far more recent public health crisis with a similarly global footprint. In September 1968, the US was confronted with a novel H3N2 influenza virus that originated in China and was dubbed the ‘Hong Kong Flu’ or ‘Mao Flu’.^1^ The death toll of the ensuing pandemic was comparable to that of the COVID-19 pandemic given the smaller US population of the time of around 200 million people, with 100,000 deaths in the US (compared to 400,000 COVID-19 deaths out of 330 million people as of January 2021). Globally, the 1968 flu pandemic resulted in between 1 to 4 million deaths. Paralleling the present day, the flu reached the White House, with both President Lyndon B. Johnson and Vice President Hubert Humphrey falling ill (Honigsbaum 2020).

Approximately one third of the US population today was alive during the 1968 pandemic (American Community Survey 2019). As shown in the map in Figure 1, the 1968 flu spread nationwide. Some communities were hit especially hard; a contemporaneous report estimated that over 40% of the population of Milwaukee was infected (Piraino et al. 1970). Several cities reported stress on local hospital systems attempting to manage the influx of patients (Saunders-Hastings and Krewski 2016; Piraino et al. 1970). Unlike SARS-CoV-2 today, the 1968 virus killed many young people, with approximately 40% of the flu-related deaths estimated to have been among those under 65 (Simonsen et al. 1998; Acosta et al. 2019).

**Figure 1:**
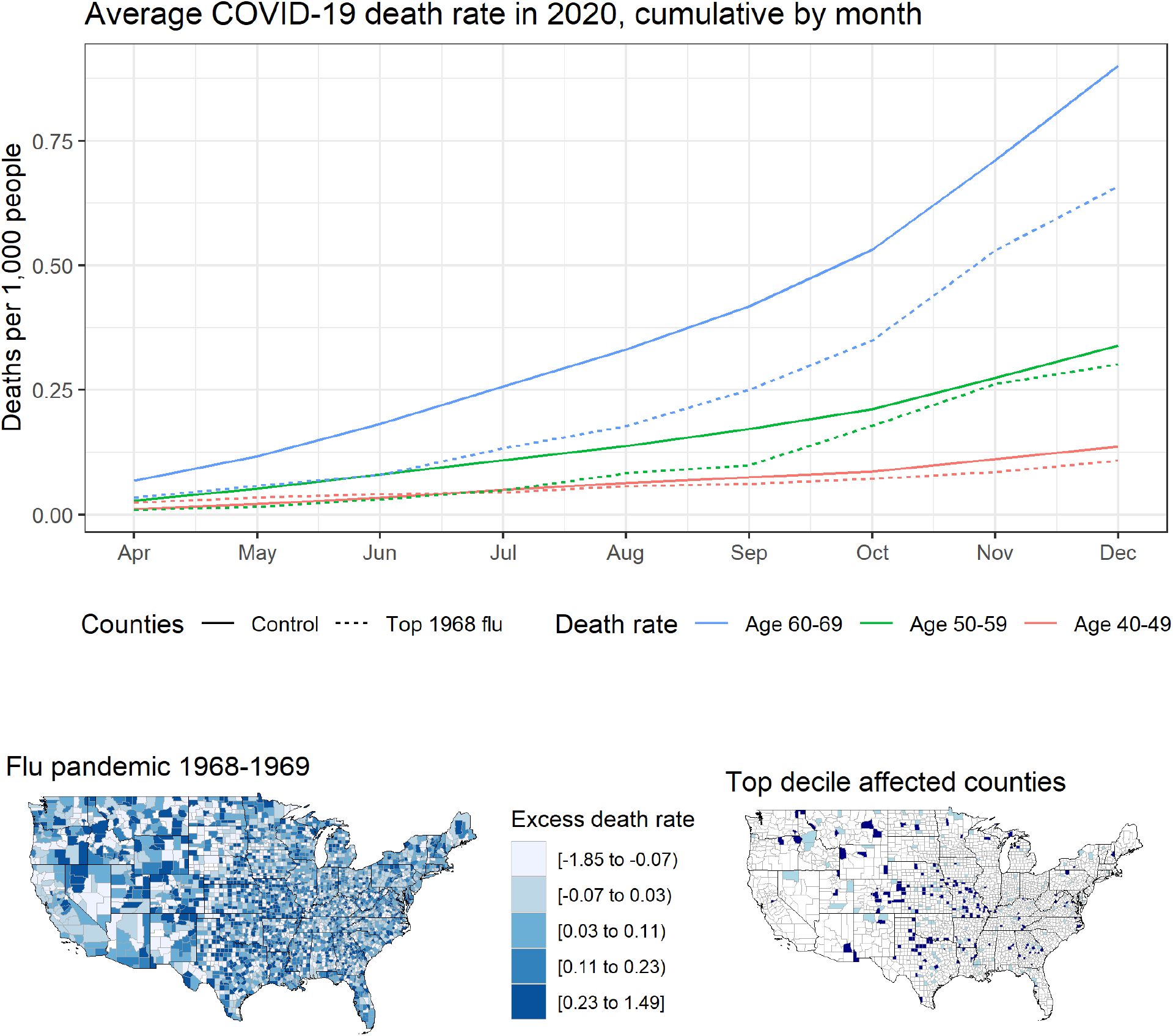
(**Top**) Average county-level cumulative COVID-19 death rates by age group and month. ‘Control’ excludes counties in the top 10% of excess flu deaths in 1968. (**Bottom**) Map of distribution of influenza deaths per thousand at the county level estimated from CDC data of excess respiratory deaths in 1968 and 1969 relative to a baseline of 1970 and 1971 (right) and a map of counties in the top 5^th^ percentile in dark blue and top 5-10^th^ percentile in light blue (right).

Despite the widespread disruptions caused by the 1968 flu pandemic, the social, economic, and public health response at the federal level was somewhat muted, with much of the country operating as usual. Although 23 states underwent school and university closures, the US did not implement any broad social distancing or containment measures. Vaccines were eventually developed but not in time to blunt the initial spread of the virus (Honigsbaum 2020; Jester et al. 2020; Saunders-Hastings and Krewski 2016).

The recency and scale of the 1968 pandemic make it a compelling event to employ historical analysis of the type seen in the third strand of literature cited above. Studying any enduring impact of this pandemic helps to further our understanding of the COVID-19 pandemic and how future pandemics can be mitigated. More immediately, it may motivate clinical research into immunological or biological responses to SARS-CoV-2 related to exposure to the 1968 pandemic.

### Empirical approach

Our overall approach is as follows: we first test the hypothesis that a residual link exists between 1968 flu severity and COVID-19 outcomes. We then assess potential factors that could explain such a relationship. While identifying precise mechanisms is outside the scope of this study, we perform several empirical tests to untangle potential *policy, social*, and *individual* channels of association. We limit the analysis to the end of 2020 to avoid potential confounding effects of differential vaccine uptake.

A *policy* channel includes any mechanism whereby an institution, in response to the 1968 flu pandemic, has put in place some deliberate action or policy that would have, regardless of its intention, influenced COVID-19 outcomes. Examples could be improved mortality outcomes as a result of hospital investments in response to the 1968 flu.

A *social* channel involves widespread behavioral shifts (e.g., in social distancing behaviors) in places that experienced high death rates from the 1968 flu, perhaps resulting in the development of a culture favoring extra precaution or social (dis)trust. In such cases, we would expect an equivalent shift for all people living within a geography (i.e., a population-level shift), as all residents would be more or less equally affected by these social forces.

An *individual* channel, in contrast, involves not a population but rather individuals for whom a difference in outcomes is expressed. The individual channel may be biological^2^ (e.g., learned immunity to the SARS-CoV-2 through prior exposure to another virus) or behavioral (e.g., individual-level compliance with public health measures), both of which may lessen the likelihood of infection or death. Along this line, Cheemarla et al. finds that previous exposure to rhinovirus lends resistance to SARS-CoV-2 infection. The individual channel requires heterogeneous effects across a population, with sub-populations demonstrating unique COVID-19 outcomes based on past exposure.

Our baseline linear regression model takes the following form:

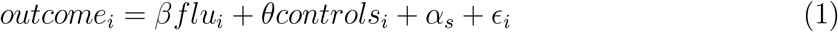

where *outcome*_*i*_ is a COVID-19 outcome in county *i* at a given month, *β* is the coefficient of interest related to *flu*, which is the excess respiratory death rate attributable to the 1968 flu as described in the Data section, *controls*_*i*_ is a vector of county-level covariates, *α*_*s*_ is a dummy for fixed effects in state *s*, and _*i*_ is the error term. Standard errors are clustered at the state level.

For outcomes of interest, we variously use (1) COVID-19 death rates, (2) hospital admissions, (3) a subset of patient-level data from Healthjump, and (4) nursing home death rates. All values are aggregated to the county level to match our data on 1968 flu intensity. We replicate this cross-sectional analysis at different snapshots in time representing the progression of the pandemic, as categorized by end-of-month outcomes.

To address endogeneity and omitted variable bias concerns in the relationship between the 1968 flu and COVID-19 outcomes, we also employ differences in death rates and hospital admissions by age group as the outcome variable. We choose the age cutoff based on whether someone was born before or after 1968 in an attempt to isolate those people likely to have been exposed to the 1968 pandemic.

The bottom panel of Figure 4 displays the identifying variation based on the age categorizations used in CDC’s COVID-19 case surveillance data and HHS’s hospital admissions data. This specification acts effectively as a difference-in-difference model to isolate the extent to which the 1968 flu affects people born before 1968 *relative* to those born after 1968. The identifying assumption relies on the fact that potential confounders are unlikely to shift the relative degree of COVID-19 morbidity or mortality across proximate age groups within a given county. Finally, to address concerns about the coarse categorization of individuals into decadal birth cohorts, we validate the results with patient-level data from Healthjump containing annual year of birth.

## Results

First, we explore patterns in the underlying data. Figure 1 plots cumulative COVID-19 death rates over time, averaged by age cohort and whether the county was among those severely hit by the 1968 flu pandemic. We see that the COVID-19 death rates are similar for the 40-49 and 50-59 groups, while the death rate among those aged 60-69, a group more likely to have been exposed to the 1968 flu, is consistently lower in high-1968-flu counties—providing suggestive evidence for our main finding.

We next present regression results using the model specified in Equation 1. Figure 2 plots the coefficients representing the change in county-level COVID-19 death rates on a cumulative basis in a given by month associated with an increase in 1968 flu mortality. We find a consistent negative relationship between the severity of 1968 outcomes and COVID-19 outcomes. Appendix Table A1 provides the full regression results including all the covariates, while Figure A1 employs various treatment dummy variables for counties greatly affected by the 1968 flu rather than a continuous measure of excess death rates. In terms of magnitude, people in counties among the 5% worst hit by the 1968 flu had COVID-19 death rates 1-4% lower than the average US county.^3^

**Figure 2:**
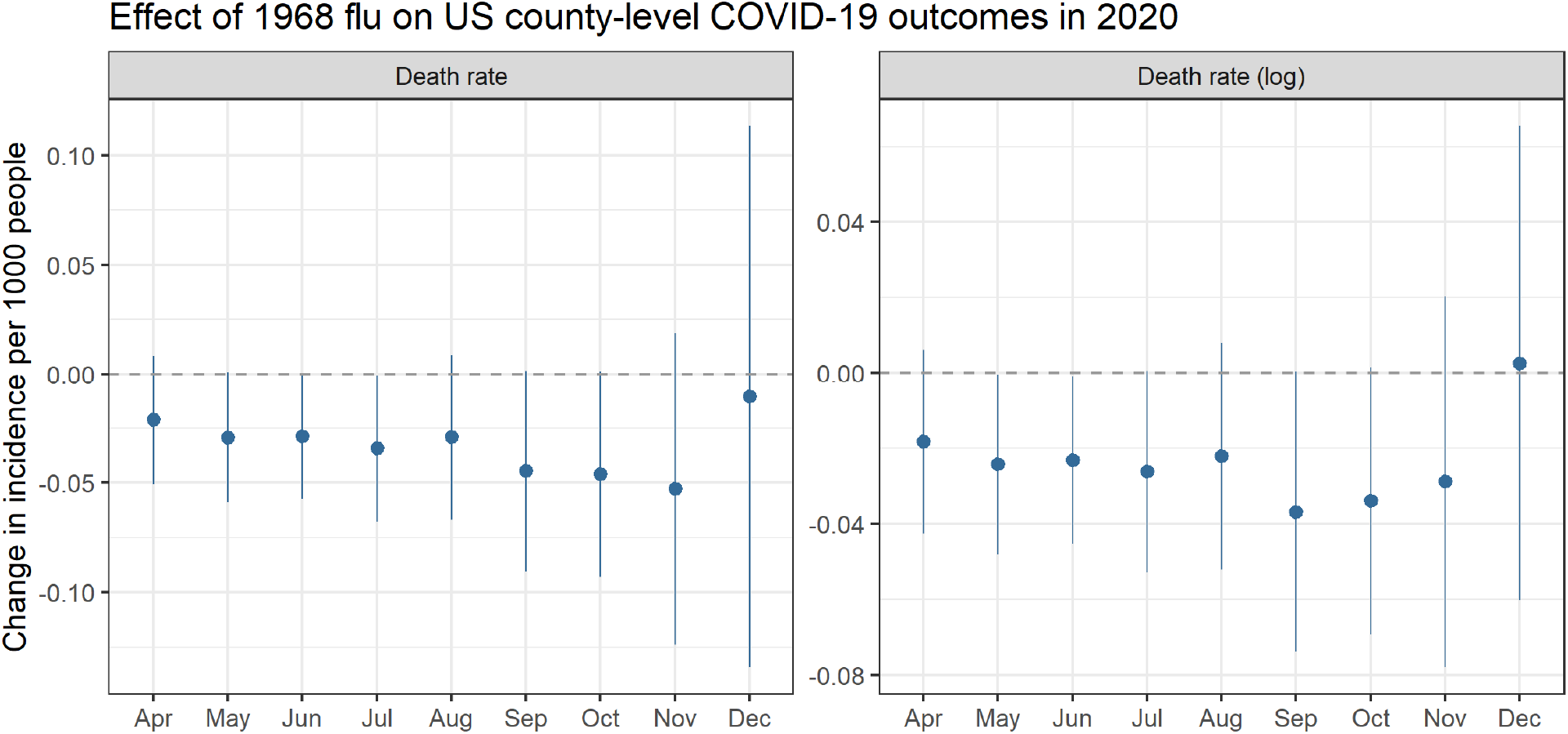
Relationship between county-level excess death rate per 1,000 from the 1968 pandemic and COVID-19 death rates (left) and log-transformed death rates (right), which can roughly be interpreted as percent change. For example, the September point estimate of −0.04 indicates a 4% reduction. All models control for socioeconomic variables and state-level fixed effects. Coefficients are plotted from separately run regressions for COVID-19 outcomes by month. Analysis ends in December 2020 at which point vaccines started to become available. Error bars reflect a 95% confidence interval.

To evaluate the hypothesis of a connection between being alive during the 1968 flu and variance in susceptibility to COVID-19, we next employ patient-level data from Healthjump containing patients’ birth year. This analysis involves 48,000 unique patient records where a COVID-19 diagnosis is explicitly linked to a medical procedure within 30 days of the diagnosis. Such procedures include, but are not limited to, hospitalization. Figure 3 plots the ratio of patients in top 10% 1968 flu counties relative to the count of all patients. Older cohorts (people alive by the 1968 pandemic) are better off in places hit hard by the 1968 flu, whereas younger cohorts are in fact *worse* off. A discontinuity appears to occur at the 1968 birth year, indeed suggesting some relationship between a likely individual experience of the 1968 flu pandemic and COVID-19 outcomes.

**Figure 3:**
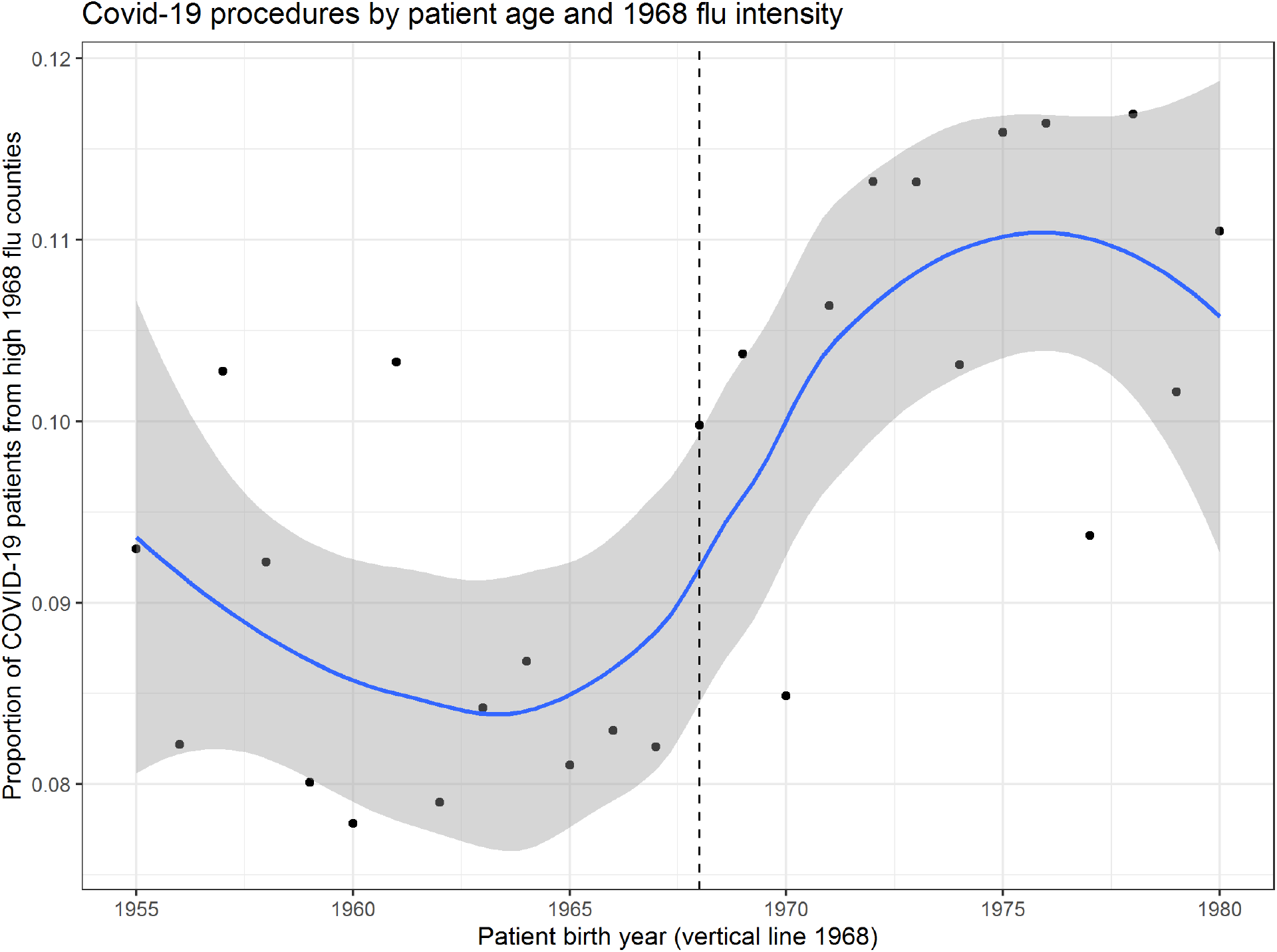
Plot of the ratio of patients in top decile 1968 flu counties relative to the count of all patients in the Healthjump dataset by patient year of birth. LOESS fit line shaded area representing a 95% confidence interval.

**Figure 4:**
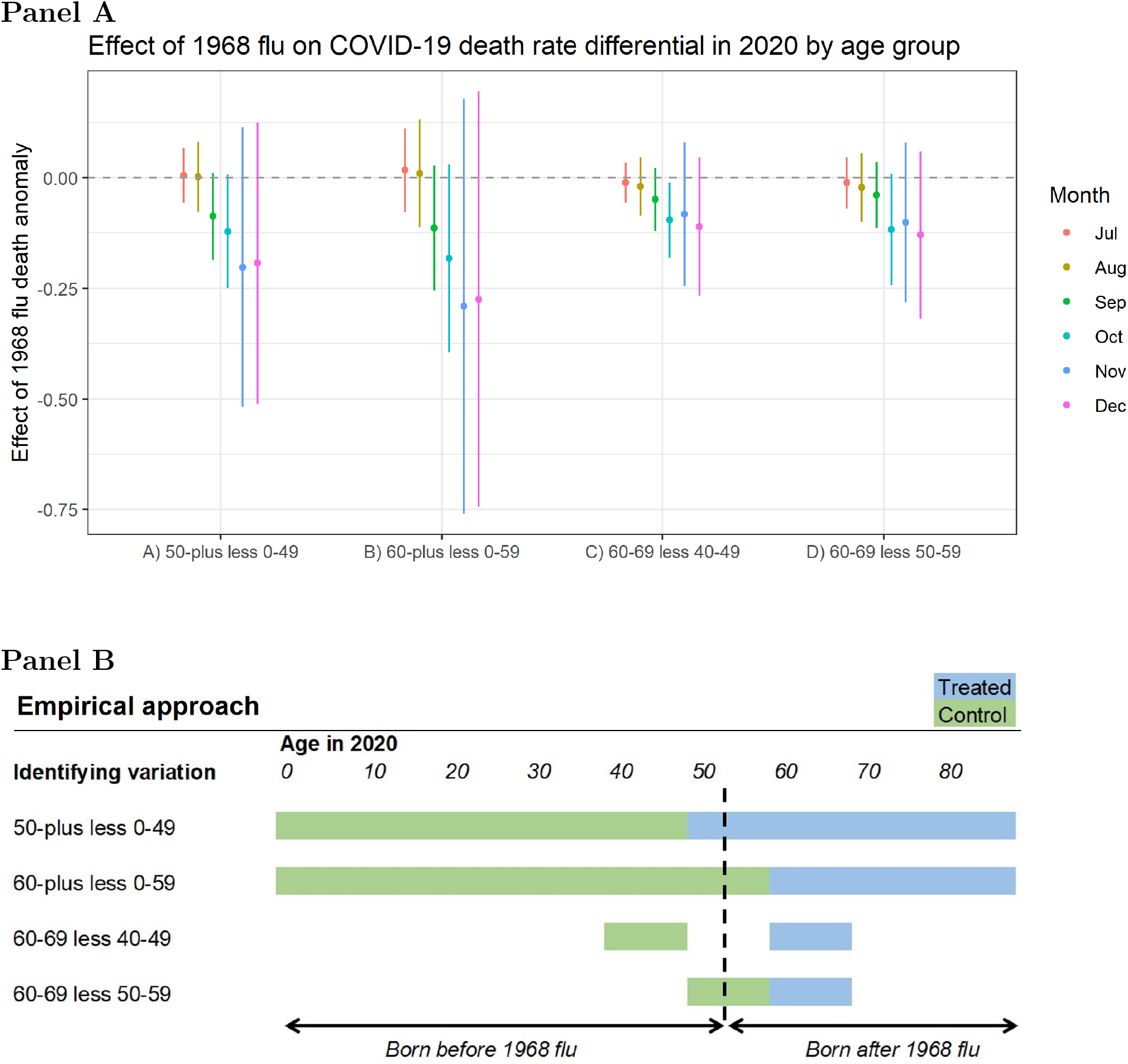
**Panel A:** Differences in county-level COVID-19 death rates per 1,000 across age groups associated with an increase in excess death rates from the 1968 pandemic, controlling for socioeconomic variables and state-level fixed effects. Coefficients are plotted from separately-run regressions for COVID-19 outcomes by month. Error bars reflect a 95% confidence interval. **Panel B:** Conceptual chart for the identifying variation used in the analysis. In the first row ‘50-plus less 0-49’ denotes the difference in the average COVID-19 death rate for the population age 50 or above (blue area) less the average death rate for the population under age 50 (green area). Multiple cutoffs are used given that the data is aggregated in decadal age brackets that do not align with 1968 pandemic exposure. The dotted line shows the theoretical age of someone in 2020 who was born in 1968.

Next, we address concerns that demographic patterns may be driving these results. We test how health outcomes relate to known confounders—both in the present and in 1968.

Appendix Table A2 presents the relationship of death rates from both COVID-19 and the 1968 flu on a selection of current and historical covariates. Models (1) and (2) show results broadly in line with the COVID-19 literature: death rates are weakly correlated with income (negative), density (negative), the elderly population (positive), and Black population (positive). However, models (3) and (4) show that present-day risk characteristics are not correlated with outcome in 1968, which reduces concerns around confounding variables. Models (5) and (6) regress 1968 flu outcomes on historical covariates. We see that the size of the elderly population is a strong predictor of death rates, supporting past literature (Acosta et al. 2019; Simonsen et al. 1998). We also note a positive, but much weaker, relationship with a county’s Black population. This finding indicates that the 1968 flu had differential impacts by race in line with the disproportionate toll of COVID-19 on the Black community (Dyer 2020), as well as Hispanics and Native Americans (Tai et al. 2020).

Appendix Figure A2 plots the mean values of the present-day covariates employed in our baseline model. Means are separately computed for top-decile counties in terms of both 1968 flu mortality and 2020 COVID-19 mortality. Values are normalized relative to a nationwide mean of 0. We see a broad correlation in average county characteristics. While not statistically different, there is gap in the average racial and ethnic composition such that COVID-19 had a more negative impact on Black and Hispanic communities relatively to the 1968 flu.

### Death rates by age group

Figure 4 plots the coefficients from Equation 1 using differences in COVID-19 death rates across age groups as the outcome variable based on CDC individual case data aggregated to the county level. There is a consistently negative point estimate for each of the four methods used to construct differentials around the age cutoffs. This means that COVID-19 death rates are lower among older cohorts relative to younger ones in places with high 1968 flu mortality.

Columns (A) and (B) showcase the difference among all those older and all those younger than 50 and 60 years old, respectively. Columns (C) and (D) limit the span to single decades. For example, column (C) shows the difference in death rates between people in their sixties versus those in their forties, highlighting groups just old enough to have lived through the 1968 pandemic and those not. Column (D) uses people in their fifties as the control, although it is less obvious how to categorize this cohort, considering that many were alive in 1968 although flu transmission tends to increase after age five with the onset of schooling (Worby et al. 2015).

### Hospital admission data

In addition to examining death rates, we look at hospital admissions driven by COVID-19 as a proxy for case severity. We again test whether there is a differential effect of 1968 flu exposure on hospitalizations between people who lived through the pandemic and those who did not. The top panel of Figure 5 presents results in two ways: first, the proportion of COVID-19 hospital admissions among people over age 60, and second, the difference in the number hospitalized among those over 60 and those under 60 years old as a proportion of the population. The 1968 flu has a negative effect on hospital admissions for the over-60 group as a whole, as well as the difference in over-60 group relative to the under-60 group (i.e., the older group was hospitalized less).

**Figure 5:**
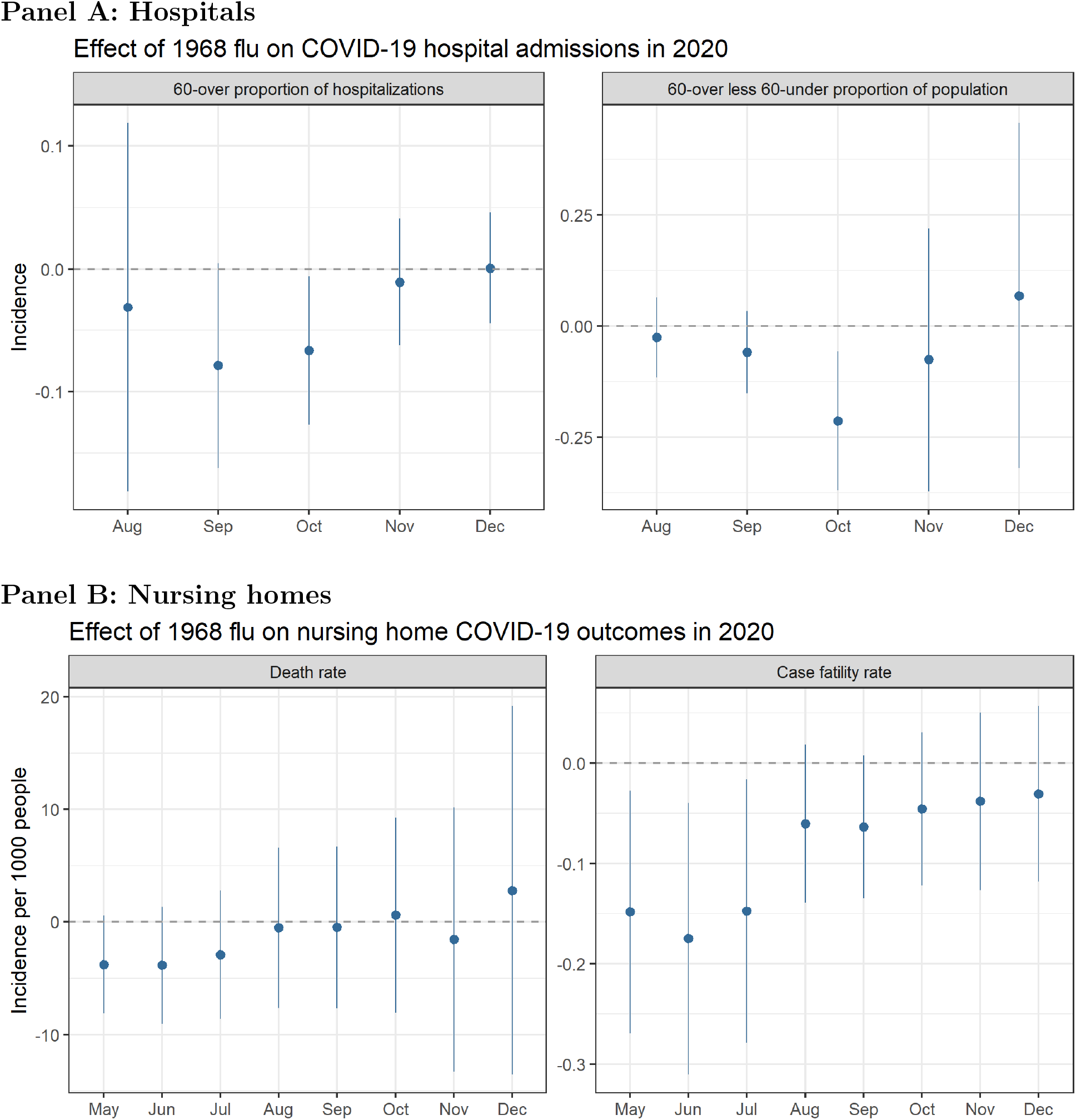
**Panel A:** Relationship between excess death rates from the 1968 pandemic and the proportion of COVID-19 hospital admissions among people over 60 (left) and the difference in the number hospitalized in the over-60 group relative to the under-60 group per 1,000 people (right). Rates are calculated based on cumulative COVID-19 hospital admissions at the county level. **Panel B:** Relationship between excess death rates from the 1968 pandemic and nursing home COVID-19 death rates, defined as cumulative deaths per 1,000 residents (left), and case fatality rate, defined as cumulative deaths as a proportion of cumulative COVID-19 cases (right). Models control for socioeconomic variables and state-level fixed effects. Coefficients are plotted from separately-run regressions for COVID-19 outcomes by month. Error bars reflect a 95% confidence interval.

### Nursing home data

We replicate our analyses using CMS data on nursing homes. The bottom panel of Figure 5 plots the effect of excess 1968 death rates on COVID-19 death rates and case fatality rates (deaths as a proportion of cases) in nursing homes. There is a negative relationship between 1968 mortality and COVID-19 mortality, but the signal is much stronger for the case fatality rate, particularly in the early months of the pandemic.

### Behavioral and institutional responses

Next, we test a number of potential relationships that would indicate potential policy, social, or individual responses to the 1968 pandemic. One potential explanation is that places with high 1968 flu mortality subsequently invested in hospital capacity to be better prepared for future challenges. Such actions could explain why hard-hit counties saw better outcomes under COVID-19. To test this, Appendix Table A3 regresses present-day number of hospital beds at the county level on 1968 flu death rates and finds no relationship—in fact, the coefficients are precise zeros under each model specification.

Another possibility is that individual behavior responds to an event like the 1968 flu. Adoption of risk-averse behaviors (whether on one’s own volition or induced through policy or social norms) could explain differential COVID-19 outcomes. Appendix Table A4 regresses a self-reported measure of mask use from the New York Times on county-level 1968 flu death rates and finds a negative relationship; that is, places worse hit by the 1968 flu tended to wear masks *less*. The relationship holds even after controlling for state-level differences and potential covariates (e.g., income, density, elderly, frontline workers).

Finally, we report changes in mobility in response to 1968 flu death rates using Google mobility data. We look at both mobility involving time spent commuting to work as well as engaging in retail and recreational activities. We compute two measures to account for the potential timing of COVID-19 responses. One is a summer average of the baseline change in mobility in a county from June to August. Another is change in county mobility between 28 days before and 28 days after a counties’ first reported death. Appendix Table A5 shows a weak but positive relationship between 1968 flu deaths and mobility, suggesting that, if anything, behavior in counties adversely affected by the 1968 flu was less compliant with directives to minimize movement than other counties. Appendix Figure A3 shows a map of mobility changes in time spent commuting to work.

## Discussion

Our findings suggest there is a persistent link between the 1968 flu pandemic and COVID-19 pandemic outcomes. People in counties among the worst hit by the 1968 flu had COVID-19 death rates 1-4% lower than the average US county. This general relationship is robust to controlling for known confounders and holds across populations (e.g., county-level aggregates, hospital admittees, and nursing home residents), as well as specifications that exploit age-based variation in exposure.

Perhaps the most salient finding in support of this conclusion is the discontinuity in Figure 3 between people born in 1968 or earlier and those born after 1968. This finding lends compelling evidence to the notion of some connection between personal experience of the 1968 flu pandemic and COVID-19 susceptibility. It is difficult to conceive of a compelling counterfactual to explain the shift in the direction of the relationship at exactly the year of the 1968 pandemic.

The direction of the relationship—that people in locales with adverse outcomes in 1968 fare *better* today—mitigates concerns of a lurking omitted variable. In such a case, we would imagine the opposite relationship in which some characteristic of places hit hard by the 1968 flu also makes them susceptible to COVID-19. Such a bias would indicate that the magnitude of our results are in fact understated.

These results suggest that the primary channel through which COVID-19 outcomes are affected is at the individual level. By contrast, we find no evidence of collective social activity suggesting better mitigation of COVID-19 outcomes. In fact, we find modestly lower levels of mask use and social distancing among the high-flu counties. Moreover, even if behaviors occurred that limited viral spread, the fact that positive outcomes were more pronounced among age cohorts that lived through the 1968 flu pandemic suggests an individual rather than societal-level mechanism.

Focusing on nursing homes also allows us to isolate a population that—unlike the elderly living outside nursing homes—exercises less agency in their social distancing practices and other risk-mitigating behaviors, instead following nursing home protocols. In addition, epidemiology within nursing homes is in some ways independent of what is found for the general population. Factors such as travel patterns of nursing home staff have a large effect on infection rates in such settings (Chen et al. 2021). We find that a smaller share of residents *who were infected* are dying. Assuming identical distributions of social distancing policy in nursing homes in counties with adverse 1968 flu histories and those without, better outcomes for nursing home residents in the former group are likely to suggest that some non-behavioral and non-policy mechanism is at play. This accumulated evidence lends support to potential biological mechanism(s) driving differential outcomes.

It is worth noting that our results are generally strongest in the autumn of 2020. The effect of the 1968 flu on COVID-19 outcomes appears to fade into the winter as COVID-19 becomes widespread, a dynamic also seen in (Lokshin et al. 2020), and as vaccines and better treatments become available. Part of this finding may be attributable to the increasing number of US deaths, which expands the number of counties making up the sample. With regard to the nursing home analysis, the more pronounced effect earlier in the COVID-19 pandemic may reflect an improvement in treatment of the disease over the course of the pandemic as standards of care within institutions evolved and more effective therapeutics were incorporated into treatment (Jorge et al. 2020).

While we cannot speak to any specific biologic mechanism, direct immunological crossreactivity seems unlikely, as SARS-CoV-2 coronavirus is a different type of virus than H3N2 influenza. However, individuals may have general innate, nonspecific, anti-viral immune pathways that are more robust due to factors such as genetics and lifestyle that may have been selected for during the 1968 pandemic and its aftermath. There is also precedent for generalized immune response experience early in life having an impact on susceptibility to future illness and injury, as has been observed in the aforementioned work examining the association between 1918 flu exposure and cardiovascular disease. Thus some sort of selective pressure may have occurred over time in which people who survived the 1968 pandemic were generally better suited to survive the COVID-19 pandemic. This mechanism would not be unlike that identified in research linking tuberculosis outcomes to the 1918 flu pandemic, and is plausible considering that 1968 flu mortality skewed toward younger people (Noymer and Garenne 2000; Simonsen et al. 1998).

These mechanisms are speculative at this point, and further research would be required to fully understand whether the differential outcomes observed here are truly meaningful and what caused them. Assessing the plausibility of cohort-based resilience to all respiratory diseases or whether this resilience is more specific to some element of COVID-19 that would otherwise cause death is certainly difficult, doing so may be worthwhile as we try to further understand this pandemic in the coming years.

### Data

County-level mortality estimates of the 1968 influenza pandemic are derived from Centers for Disease Control and Prevention (CDC) Compressed Mortality files, 1968-1978, accessed via CDC’s WONDER database (Centers for Disease Control and Prevention (CDC), National Center for Health Statistics 2000). We estimate the excess influenza death rate by comparing excess respiratory deaths in 1968 and 1969 (when the vast majority of 1968 flu deaths occur) to a baseline period of 1970 and 1971. We use death rates defined as deaths per thousand people using local population estimates at the time. Our methodology of estimating excess mortality follows previous work estimating mortality attributable to pandemic flu (Alling et al. 1981).

Individual-level data are from CDC’s COVID-19 Case Surveillance Restricted Access Detailed Data accessed January 2021. The restricted dataset includes over 12 million records of COVID-19 cases with date, decadal age group, and county identifiers. Deaths are also reported. Note that because of CDC reporting delays and state-level data filing practices, aggregate totals are less than those of other sources and records November 2020 onward contain fewer counties reporting than earlier (*COVID-19 Case Surveillance Restricted Access Data 2021*).

Hospital admission data are gathered from the US Department of Health and Housing Services (HHS). We aggregate weekly-level hospital data for different decadal age groups to the county-month level (*COVID-19 Reported Patient Impact and Hospital Capacity by Facility* 2021). Nursing home data come from the Centers for Medicare Medicaid Services (CMS) Nursing Home COVID-19 Public File. Nursing home facilities are required to self-report these data to the CDC.^4^ Patient-level healthcare data with year-of-birth information available from Healthjump via the COVID-19 Research Database consortium (COVID-19 Research Database 2021). The number of hospital beds at the county level are derived from the Centers for Medicare Medicaid Services. Mask use survey data come from the New York Times (Katz et al. 2020). County-level mobility data were made accessible to COVID-19 researchers by Google (Google 2020).

Covariate data include county-level race, ethnicity and age structure data from the US Census and mean county-level income data from the US Bureau of Economic Analysis (SEER Program, National Cancer Institute, NIH 2020; US Bureau of Economic Analysis 2020). Data on nursing home populations, incarcerated populations, uninsured populations, average household size, and work commuting methods come from the 2014-2018 American Community Survey (U.S. Census Bureau 2019c; 2019e; 2019b; 2019d). Data on manufacturing establishments come from the American Economic Survey (2019a). Number of frontline workers were derived from CEPR data (Fremstad et al. 2020), transforming to the county level assuming even allocation. The freight index is from the FHA’s Freight Analysis Framework (U.S. Department of Transportation 2020).

## Data Availability

All data except for the Healthjump data produced in the present study are available upon reasonable request to the authors. Restrictions apply to the availability of the Healthjump data, which were used under license by the COVID-19 Research Database consortium for the current study, and so are not publicly available. Healthjump data are available with permission of the COVID-19 Research Database consortium.

## Data Availability

Data used for the purpose of this study will be made accessible upon publishing at https://www.github.com/cboulos/Flu-1968

## Appendix

### Figures

**Figure A1:**
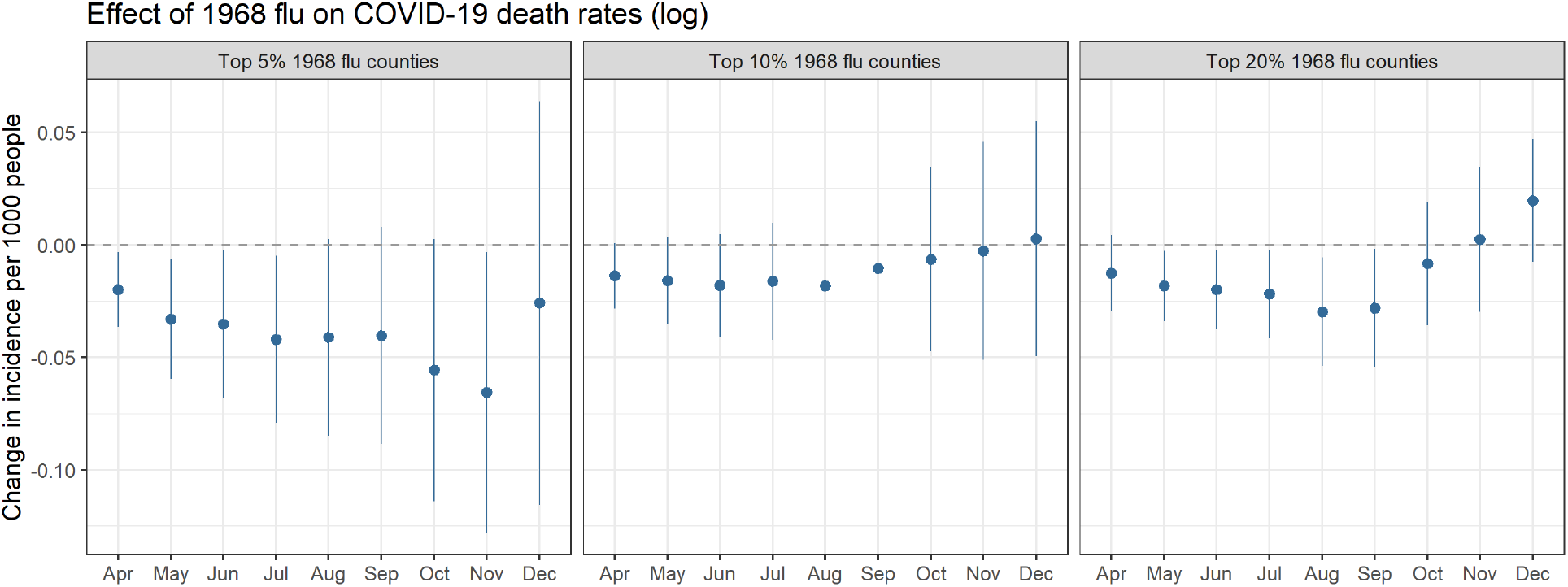
Relationship between cumulative log-transformed COVID-19 death rates at the county level by month and an indicator if counties are in the top 5^th^ (**left**), 10^th^ (**center**), or 20^th^ (**right**) percentile of excess 1968 pandemic death rates, controlling for socioeconomic variables and state-level fixed effects. Coefficients are plotted from separately-run regressions for COVID-19 outcomes by month. Error bars reflect a 95% confidence interval.

**Figure A2:**
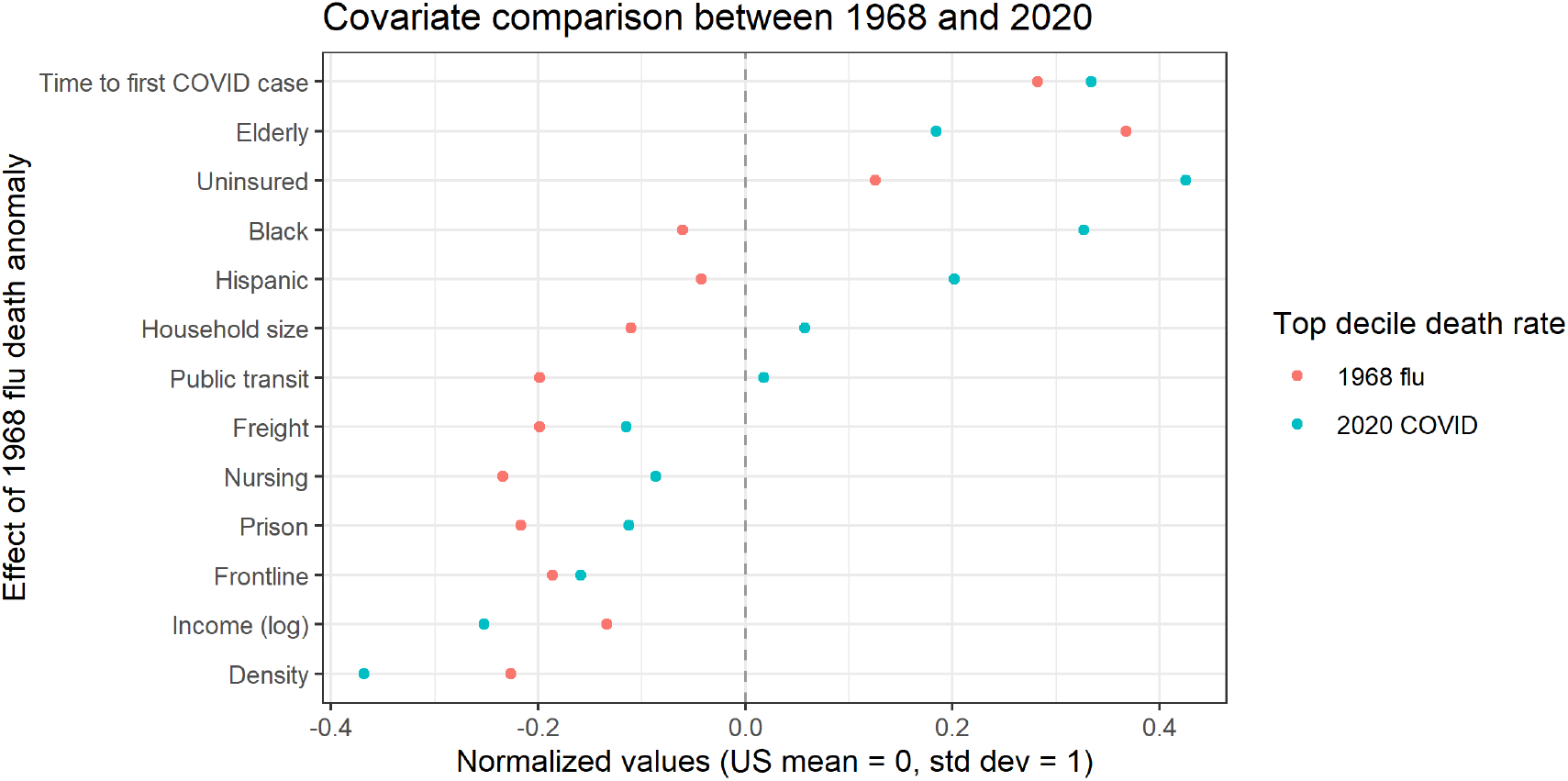
Plot of the mean values of the present-day covariates utilized in our baseline model. Means are separately computed for top decile counties in terms of both 1968 flu mortality and 2020 COVID-19 mortality. Values are normalized relative to a nation-wide mean of 0. We see a broad correlation in average county characteristics.

**Figure A3:**
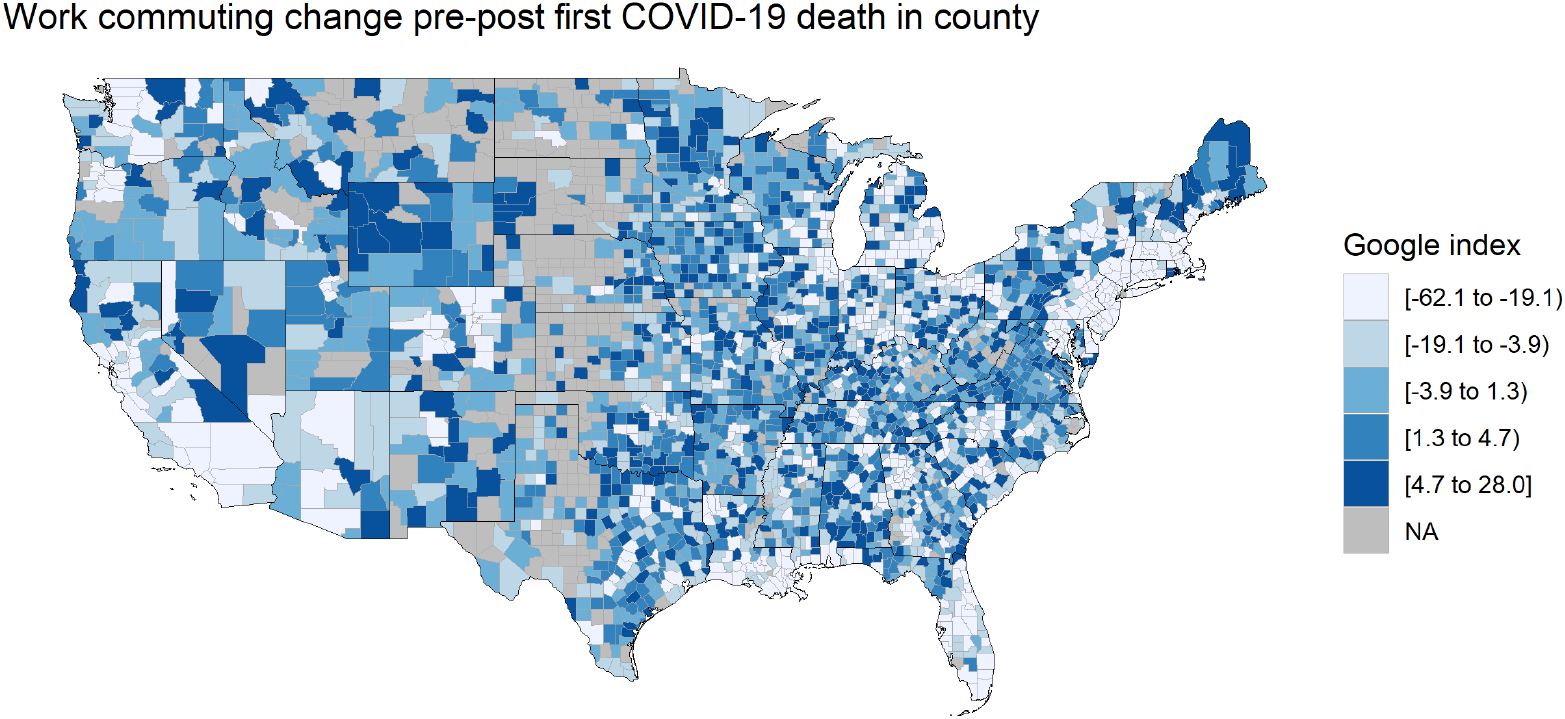
County-level map of Google mobility data on changes in time spent commuting to work 28 days before and after the first county death.

### Tables

**Table A1:** Impact of 1968 flu on county COVID-19 deaths, cumulative by month

|  | Deaths per 1,000 as of month |  |  |  |  |  |  |
| --- | --- | --- | --- | --- | --- | --- | --- |
|  | Jun | Jul | Aug | Sep | Oct | Nov | Dec |
|  | (1) | (2) | (3) | (4) | (5) | (6) | (7) |
| Flu deaths 1968 | −0.029*<br>(0.015) | −0.034*<br>(0.017) | −0.029<br>(0.019) | −0.045*<br>(0.023) | −0.046*<br>(0.024) | −0.053<br>(0.036) | −0.010<br>(0.063) |
| Income per capita (log) | 0.039<br>(0.031) | 0.016<br>(0.030) | −0.053<br>(0.033) | −0.088**<br>(0.042) | −0.152**<br>(0.059) | −0.269***<br>(0.073) | −0.369***<br>(0.097) |
| Density | 0.033<br>(0.022) | 0.027<br>(0.024) | 0.023<br>(0.026) | 0.010<br>(0.026) | −0.009<br>(0.033) | −0.032<br>(0.041) | −0.122**<br>(0.053) |
| Density-squared | −0.006<br>(0.005) | −0.005<br>(0.005) | −0.005<br>(0.005) | −0.004<br>(0.005) | −0.002<br>(0.006) | −0.001<br>(0.007) | 0.007<br>(0.009) |
| Timing first case | −0.285***<br>(0.064) | −0.349***<br>(0.070) | −0.332***<br>(0.091) | −0.327***<br>(0.114) | −0.255**<br>(0.115) | −0.152<br>(0.210) | −0.187<br>(0.250) |
| Elderly proportion | 1.053***<br>(0.285) | 1.302***<br>(0.270) | 1.726***<br>(0.311) | 2.015***<br>(0.397) | 2.419***<br>(0.483) | 2.970***<br>(0.640) | 3.855***<br>(0.927) |
| Black proportion | 0.846***<br>(0.172) | 1.014***<br>(0.173) | 1.176***<br>(0.170) | 1.293***<br>(0.152) | 1.289***<br>(0.147) | 1.132***<br>(0.145) | 0.865***<br>(0.181) |
| Hispanic proportion | 0.139<br>(0.115) | 0.347**<br>(0.133) | 0.660***<br>(0.204) | 0.809***<br>(0.279) | 0.884**<br>(0.329) | 0.971**<br>(0.372) | 1.075***<br>(0.385) |
| Freight intensity | 0.007<br>(0.009) | 0.009<br>(0.011) | 0.009<br>(0.012) | 0.010<br>(0.014) | 0.011<br>(0.014) | 0.015<br>(0.017) | 0.023<br>(0.018) |
| Public transit proportion | 0.016**<br>(0.007) | 0.015**<br>(0.007) | 0.014**<br>(0.006) | 0.012*<br>(0.006) | 0.014**<br>(0.006) | 0.017**<br>(0.007) | 0.015<br>(0.009) |
| Household size | 0.138***<br>(0.051) | 0.188***<br>(0.056) | 0.199***<br>(0.063) | 0.209***<br>(0.066) | 0.189**<br>(0.075) | 0.077<br>(0.103) | −0.041<br>(0.141) |
| Nursing home proportion | 0.00005***<br>(0.00001) | 0.0001***<br>(0.00001) | 0.0001***<br>(0.00001) | 0.0001***<br>(0.00001) | 0.0001***<br>(0.00001) | 0.00005***<br>(0.00002) | 0.00004**<br>(0.00002) |
| Prisoner proportion | −0.00001*<br>(0.00001) | −0.00001*<br>(0.00001) | −0.00001*<br>(0.00001) | −0.00001*<br>(0.00001) | −0.00001<br>(0.00001) | −0.00001<br>(0.00001) | −0.00001<br>(0.00001) |
| Uninsured proportion | 0.002<br>(0.002) | 0.004<br>(0.003) | 0.005*<br>(0.003) | 0.007**<br>(0.003) | 0.010**<br>(0.005) | 0.016***<br>(0.006) | 0.021**<br>(0.008) |
| Frontline proportion | −0.00000***<br>(0.00000) | −0.00000***<br>(0.00000) | −0.00000***<br>(0.00000) | −0.00000***<br>(0.00000) | −0.00000***<br>(0.00000) | −0.00000***<br>(0.00000) | −0.00000**<br>(0.00000) |
| Controls | X | X | X | X | X | X | X |
| State FE | X | X | X | X | X | X | X |
| Observations | 3,024 | 3,024 | 3,024 | 3,024 | 3,024 | 3,024 | 3,024 |
| R<sup>2</sup> | 0.416 | 0.437 | 0.478 | 0.464 | 0.382 | 0.335 | 0.331 |

Notes: Linear regression. Standard errors clustered at state level. \*p<0.1; \*\*p<0.05; \*\*\*p<0.01

**Table A2:** Impact of current and past characteristics on COVID-19 and 1968 flu death rates

|  | *Dependent variable:* |  |  |  |  |  |
| --- | --- | --- | --- | --- | --- | --- |
|  | COVID death rate |  | 1968 flu death rate |  |  |  |
|  | –Current covariates– |  | –Current covariates– |  | –1969 covariates– |  |
|  | (1) | (2) | (3) | (4) | (5) | (6) |
| Income per capita (log) | –0.193<br>(0.140) | –0.372***<br>(0.095) | –0.024<br>(0.032) | 0.002<br>(0.029) | 0.022<br>(0.031) | 0.044<br>(0.034) |
| Density | 0.026<br>(0.106) | 0.079<br>(0.055) | 0.002<br>(0.009) | –0.002<br>(0.010) | 0.001<br>(0.007) | 0.001<br>(0.007) |
| Elderly proportion | 1.333*<br>(0.759) | 1.549**<br>(0.682) | –0.006<br>(0.182) | 0.008<br>(0.192) | 0.957***<br>(0.266) | 1.099***<br>(0.245) |
| Black proportion | 1.527***<br>(0.299) | 1.070***<br>(0.207) | 0.047<br>(0.047) | 0.014<br>(0.052) | 0.124**<br>(0.050) | 0.059<br>(0.067) |
| State FE |  | X |  | X |  | X |
| Observations | 3,007 | 3,007 | 3,007 | 3,007 | 3,007 | 3,007 |
| R<sup>2</sup> | 0.109 | 0.298 | 0.001 | 0.030 | 0.012 | 0.039 |

*Notes:* Linear regression. COVID-19 deaths as of Nov 2020. We exclude covariates for which we lack data in 1968 to ensure similar sample composition. Standard errors clustered at state level. \*p<0.1; \*\*p<0.05; \*\*\*p<0.01

**Table A3:**
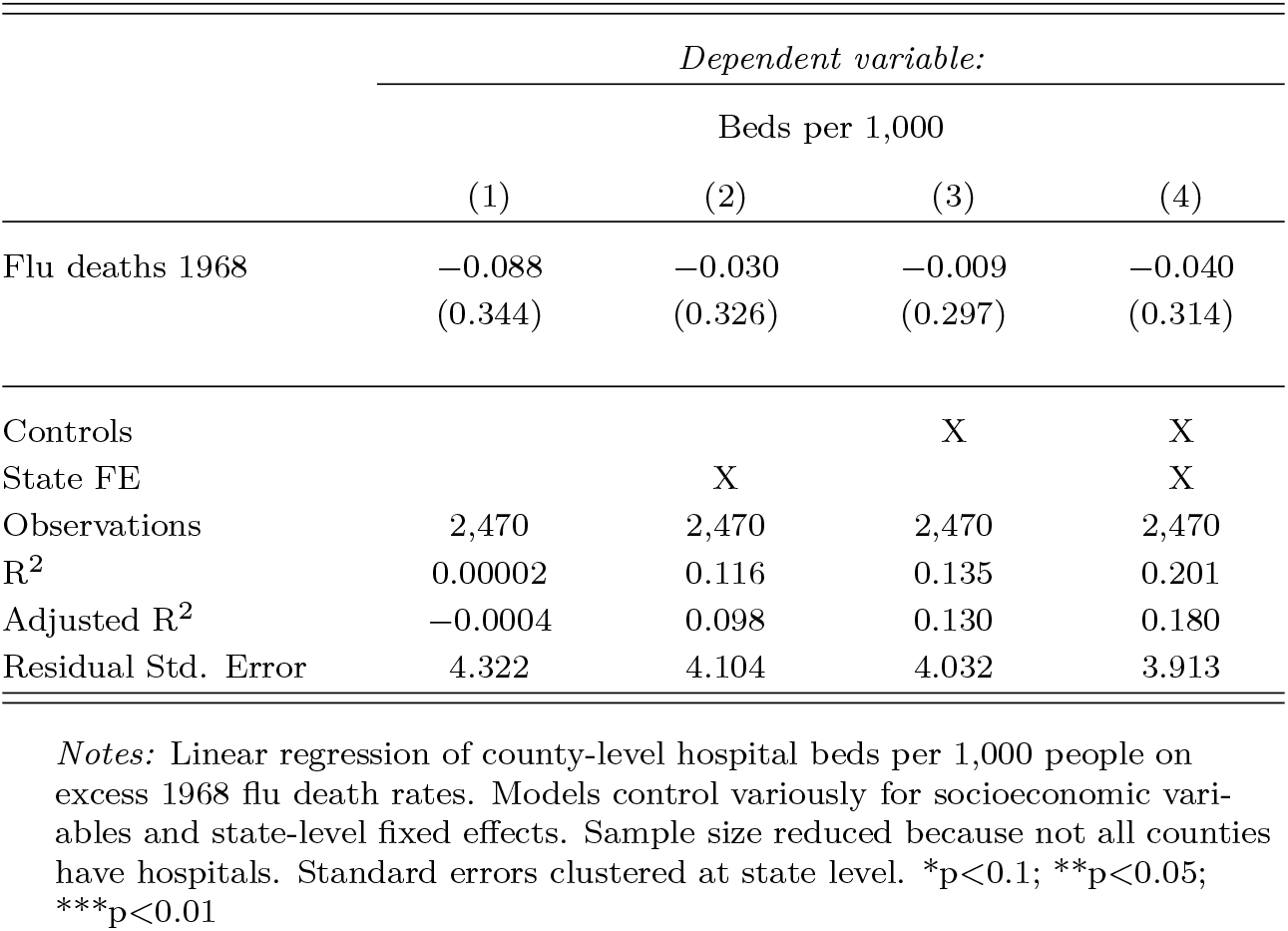
Impact of 1968 flu on current number of hospital beds

|  | *Dependent variable:* |  |  |  |
| --- | --- | --- | --- | --- |
|  | Beds per 1,000 |  |  |  |
|  | (1) | (2) | (3) | (4) |
| Flu deaths 1968 | −0.088<br>(0.344) | −0.030<br>(0.326) | −0.009<br>(0.297) | −0.040<br>(0.314) |
| Controls |  |  | X | X |
| State FE |  | X |  | X |
| Observations | 2,470 | 2,470 | 2,470 | 2,470 |
| R<sup>2</sup> | 0.00002 | 0.116 | 0.135 | 0.201 |
| Adjusted R<sup>2</sup> | −0.0004 | 0.098 | 0.130 | 0.180 |
| Residual Std. Error | 4.322 | 4.104 | 4.032 | 3.913 |

*Notes:* Linear regression of county-level hospital beds per 1,000 people on excess 1968 flu death rates. Models control variously for socioeconomic variables and state-level fixed effects. Sample size reduced because not all counties have hospitals. Standard errors clustered at state level. \*p<0.1; \*\*p<0.05; \*\*\*p<0.01

**Table A4:** Impact of 1968 flu on reported mask use frequency

|  | *Dependent variable:* |  |  |  |
| --- | --- | --- | --- | --- |
|  | Mask use propensity |  |  |  |
|  | (1) | (2) | (3) | (4) |
| Flu deaths 1968 | −0.017**<br>(0.008) | −0.012**<br>(0.005) | −0.013**<br>(0.005) | −0.011**<br>(0.005) |
| Controls |  |  | X | X |
| State FE |  | X |  | X |
| Observations | 3,024 | 3,024 | 3,024 | 3,024 |
| R<sup>2</sup> | 0.002 | 0.374 | 0.248 | 0.419 |
| Adjusted R<sup>2</sup> | 0.002 | 0.364 | 0.244 | 0.407 |
| Residual Std. Error | 0.088 | 0.070 | 0.077 | 0.068 |

*Notes:* Linear regression of county-level mask use defined as proportion of sample responding to wearing masks ‘Sometimes’ or ‘Frequently’. Models control variously for socioeconomic variables and state-level fixed effects. Standard errors clustered at state level \*p<0.1; \*\*p<0.05; \*\*\*p<0.01

**Table A5:**
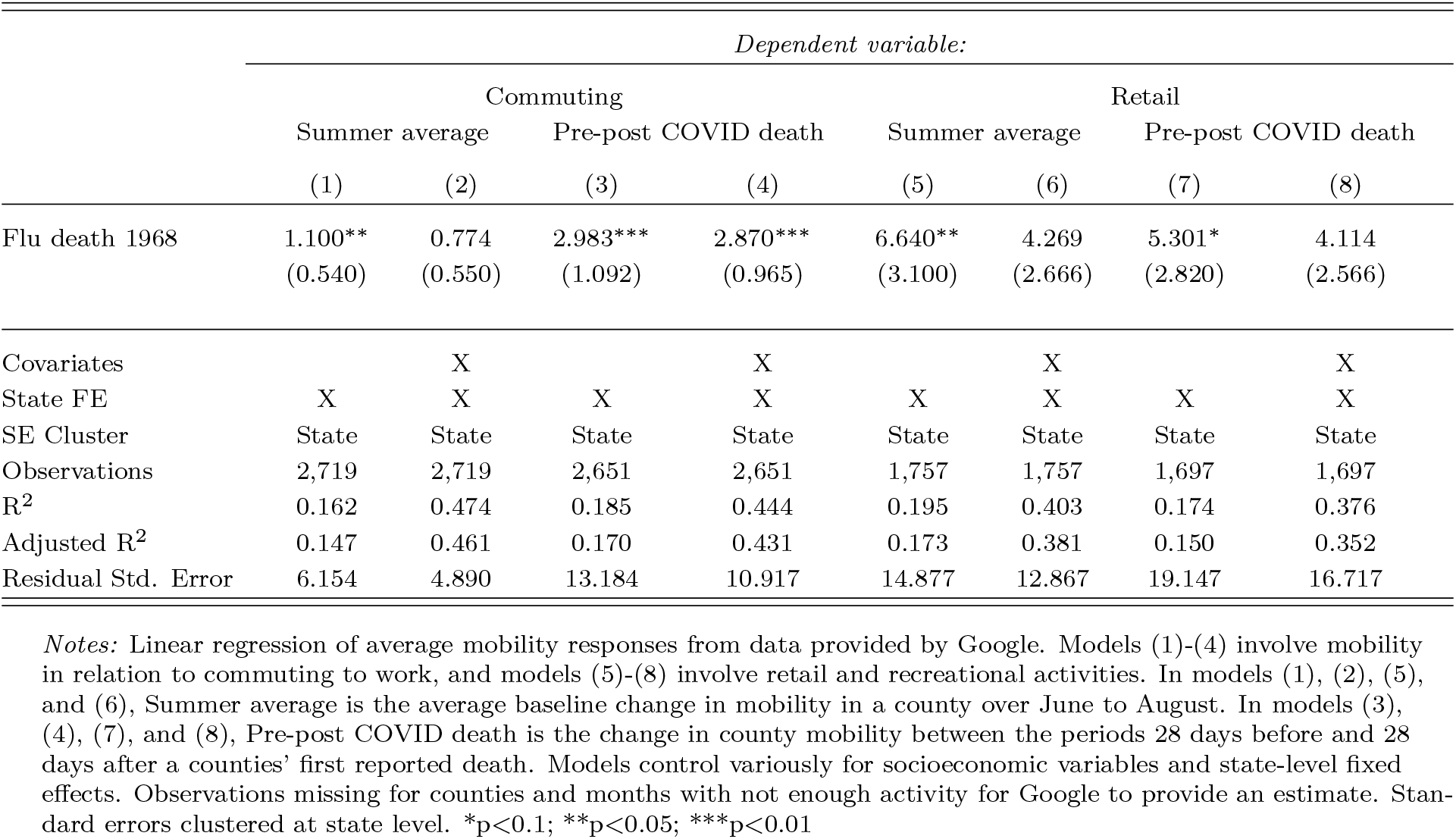
Impact of 1968 pandemic on mobility anomalies from Google

|  | *Dependent variable:* |  |  |  |  |  |  |  |
| --- | --- | --- | --- | --- | --- | --- | --- | --- |
|  | Commuting |  |  |  | Retail |  |  |  |
|  | Summer average |  | Pre-post COVID death |  | Summer average |  | Pre-post COVID death |  |
|  | (1) | (2) | (3) | (4) | (5) | (6) | (7) | (8) |
| Flu death 1968 | 1.100**<br>(0.540) | 0.774<br>(0.550) | 2.983***<br>(1.092) | 2.870***<br>(0.965) | 6.640**<br>(3.100) | 4.269<br>(2.666) | 5.301*<br>(2.820) | 4.114<br>(2.566) |
| Covariates |  | X |  | X |  | X |  | X |
| State FE | X | X | X | X | X | X | X | X |
| SE Cluster | State | State | State | State | State | State | State | State |
| Observations | 2,719 | 2,719 | 2,651 | 2,651 | 1,757 | 1,757 | 1,697 | 1,697 |
| R<sup>2</sup> | 0.162 | 0.474 | 0.185 | 0.444 | 0.195 | 0.403 | 0.174 | 0.376 |
| Adjusted R<sup>2</sup> | 0.147 | 0.461 | 0.170 | 0.431 | 0.173 | 0.381 | 0.150 | 0.352 |
| Residual Std. Error | 6.154 | 4.890 | 13.184 | 10.917 | 14.877 | 12.867 | 19.147 | 16.717 |

*Notes:* Linear regression of average mobility responses from data provided by Google. Models (1)-(4) involve mobility in relation to commuting to work, and models (5)-(8) involve retail and recreational activities. In models (1), (2), (5), and (6), Summer average is the average baseline change in mobility in a county over June to August. In models (3), (4), (7), and (8), Pre-post COVID death is the change in county mobility between the periods 28 days before and 28 days after a counties' first reported death. Models control variously for socioeconomic variables and state-level fixed effects. Observations missing for counties and months with not enough activity for Google to provide an estimate. Standard errors clustered at state level. \*p<0.1; \*\*p<0.05; \*\*\*p<0.01

## Footnotes

1 https://www.cdc.gov/flu/pandemic-resources/1968-pandemic.html

2 An example of such a channel is how influenza affects age groups differently based on individuals’ past exposure to a similar viruses (Acosta et al. 2019; Reichert et al. 2012; Ma et al. 2011). Further, prenatal exposure to the 1918 influenza virus has been linked to cardiovascular disease and other phenomena occurring decades later (Almond 2006; Mazumder et al. 2010), although others have disputed some of these links (Beach et al. 2022).

3 Magnitude range is computed in two ways: first, using the continuous measure in Figure 2, top 5% counties had an average 1968 flu death rate of 0.67 per thousand compared to the US average of 0.07, for a difference of 0.6 deaths per thousand, or almost 10 times the death rate. Multiplying 0.6 by the average of the June to December coefficients from the right panel (log-value) of Figure 2 (−0.023) equals −0.014, or −1.4%. Alternatively, in the model from Figure A1, employing a binary treatment for top 5% 1968 flu counties, the average of the coefficients is −0.04, or −4%. Putting the two approaches together results in a reduction range of 1-4%.

4 While CDC successfully consolidated data from nursing homes across the US, there have been reports that the data received may not be comprehensive (Attorney General 2021; *COVID-19 Nursing Home Data* 2021).

## References

Abildgren, Kim. 2021. “Archival Big Data and the Spanish Flu in Copenhagen.” Information Discovery and Delivery ahead-of-print (ahead-of-print). ISSN: 2398-6247. doi:10.1108/IDD-11-2020-0142. https://doi.org/10.1108/IDD-11-2020-0142.

Acosta, Enrique, Stacey A Hallman, Lisa Y Dillon, Nadine Ouellette, Robert Bourbeau, D Ann Herring, Kris Inwood, David JD Earn, Joaquin Madrenas, Matthew S Miller, et al. 2019. “Determinants of influenza mortality trends: age-period-cohort analysis of influenza mortality in the United States, 1959–2016.” Demography 56 (5): 1723–1746.

Alling, DW, WC Blackwelder, CH Stuart-harris, et al. 1981. “A study of excess mortality during influenza epidemics in the United States, 1968-1976.” American Journal of Epidemiology 113 (1): 30–43.

Almond, Douglas. 2006. “Is the 1918 influenza pandemic over? Long-term effects of in utero influenza exposure in the post-1940 US population.” Journal of political Economy 114 (4): 672–712.

American Community Survey. 2019. S0101: Age and Sex. U.S. Department of Commerce. https://data.census.gov/cedsci/table?q=population%5C%20census&tid=ACSST1Y2019.S0101&hidePreview=false.

Attorney General, New York State Office of the. 2021. Attorney General James Releases Report on Nursing Homes’ Response to COVID-19 | New York State Attorney General. https://ag.ny.gov/press-release/2021/attorney-general-james-releases-report-nursing-homes-response-covid-19. (Accessed on 01/31/2021).

Beach, Brian, Ryan Brown, Joseph Ferrie, Martin Saavedra, and Duncan Thomas. 2022. “Re-evaluating the Long-Term Impact of In Utero Exposure to the 1918 Influenza Pandemic.”

Beach, Brian, Karen Clay, and Martin H Saavedra. 2020. “The 1918 influenza pandemic and its lessons for COVID-19.” National Bureau of Economic Research Working Paper Series, no. w27673.

Centers for Disease Control and Prevention (CDC), National Center for Health Statistics. 2000. Compressed Mortality File 1968-1978. Accessed February 2, 2021. http://wonder.cdc.gov/cmf-icd8.html.

Cheemarla, Nagarjuna R., Timothy A. Watkins, Valia T. Mihaylova, Bao Wang, Dejian Zhao, Guilin Wang, Marie L. Landry, and Ellen F. Foxman. 2021. “Dynamic innate immune response determines susceptibility to SARS-CoV-2 infection and early replication kinetics.” E20210583, Journal of Experimental Medicine 218 (8). ISSN: 0022-1007. doi:10.1084/jem.20210583. https://doi.org/10.1084/jem.20210583.

Chen, M. Keith, Judith A. Chevalier, and Elisa F. Long. 2021. “Nursing home staff networks and COVID-19.” Proceedings of the National Academy of Sciences 118 (1). ISSN: 0027-8424. doi:10.1073/pnas.2015455118. eprint: https://www.pnas.org/content/118/1/e2015455118.full.pdf. https://www.pnas.org/content/118/1/e2015455118.

Cheng, Vincent Chi-Chung, Shuk-Ching Wong, Vivien Wai-Man Chuang, Simon Yung-Chun So, Jonathan Hon-Kwan Chen, Siddharth Sridhar, Kelvin Kai-Wang To, Jasper Fuk-Woo Chan, Ivan Fan-Ngai Hung, Pak-Leung Ho, et al. 2020. “The role of community-wide wearing of face mask for control of coronavirus disease 2019 (COVID-19) epidemic due to SARS-CoV-2.” Journal of Infection 81 (1): 107–114.

COVID-19 Case Surveillance Restricted Access Data. 2021. Centers for Disease Control and Prevention. Accessed June 17, 2021. https://data.cdc.gov/d/mbd7-r32t.

COVID-19 Nursing Home Data. 2021. CMS - Division of Nursing Homes/Quality, Safety, and Oversight Group/Center for Clinical Standards and Quality. Accessed June 17, 2021. https://data.cms.gov/d/bkwz-xpvg.

COVID-19 Reported Patient Impact and Hospital Capacity by Facility. 2021. Centers for Disease Control and Prevention. Accessed June 17, 2021. https://healthdata.gov/d/j4ip-wfsv.

COVID-19 Research Database. 2021. Accessed June 15, 2021. https://covid19researchdatabase.org.

Doshi, Peter. 2020. “Covid-19: Do many people have pre-existing immunity?” Bmj 370.

Dowd, Jennifer Beam, Liliana Andriano, David M Brazel, Valentina Rotondi, Per Block, Xuejie Ding, Yan Liu, and Melinda C Mills. 2020. “Demographic science aids in understanding the spread and fatality rates of COVID-19.” Proceedings of the National Academy of Sciences 117 (18): 9696–9698.

Dyer, Owen. 2020. Covid-19: Black people and other minorities are hardest hit in US.

Fremstad, Shawn, Hye Jin Rho, and Hayley Brown. 2020. “Meatpacking Workers are a Diverse Group Who Need Better Protections.” https://cepr.net/meatpacking-workers-are-a-diverse-group-who-need-better-protections/.

Galvani, Alison P, and Montgomery Slatkin. 2003. “Evaluating plague and smallpox as historical selective pressures for the CCR5-Δ32 HIV-resistance allele.” Proceedings of the National Academy of Sciences 100 (25): 15276–15279.

Google. 2020. Google COVID-19 Community Mobility Reports. https://www.google.com/covid19/mobility/. Accessed May 9, 2020.

Group, Severe Covid-19 GWAS. 2020. “Genomewide association study of severe Covid-19 with respiratory failure.” New England Journal of Medicine 383 (16): 1522–1534.

Honigsbaum, Mark. 2020. “Revisiting the 1957 and 1968 influenza pandemics.” The Lancet 395 (10240): 1824–1826.

Hou, Yuan, Junfei Zhao, William Martin, Asha Kallianpur, Mina K Chung, Lara Jehi, Nima Sharifi, Serpil Erzurum, Charis Eng, and Feixiong Cheng. 2020. “New insights into genetic susceptibility of COVID-19: an ACE2 and TMPRSS2 polymorphism analysis.” BMC medicine 18 (1): 1–8.

Huang, Junjie, Jeremy Yuen-Chun Teoh, Sunny H. Wong, and Martin C. S. Wong. 2020. “The potential impact of previous exposure to SARS or MERS on control of the COVID-19 pandemic.” European Journal of Epidemiology 35 (11): 1099–1103. ISSN: 1573-7284. doi:10.1007/s10654-020-00674-9. https://doi.org/10.1007/s10654-020-00674-9.

Jester, Barbara J, Timothy M Uyeki, and Daniel B Jernigan. 2020. “Fifty years of influenza A (H3N2) following the pandemic of 1968.” American journal of public health 110 (5): 669–676.

Jorge, April, Kristin M D’Silva, Andrew Cohen, Zachary S Wallace, Natalie McCormick, Yuqing Zhang, and Hyon K Choi. 2020. “Temporal trends in severe COVID-19 outcomes in patients with rheumatic disease: a cohort study.” The Lancet Rheumatology.

Katz, Josh, Margot Sanger-Katz, and Kevin Quealy. 2020. “A detailed map of who is wearing masks in the US.” The New York Times.

Knittel, Christopher R, and Bora Ozaltun. 2020. What Does and Does Not Correlate with COVID-19 Death Rates. Working Paper, Working Paper Series 27391. National Bureau of Economic Research. doi:10.3386/w27391. http://www.nber.org/papers/w27391.

Kraemer, Moritz UG, Chia-Hung Yang, Bernardo Gutierrez, Chieh-Hsi Wu, Brennan Klein, David M Pigott, Open COVID-19 Data Working Group †, Louis du Plessis, Nuno R Faria, Ruoran Li, et al. 2020. “The effect of human mobility and control measures on the COVID-19 epidemic in China.” Science 368 (6490): 493–497.

Lauc, Gordan, and David Sinclair. 2020. “Biomarkers of biological age as predictors of COVID-19 disease severity.” Aging (Albany NY) 12 (8): 6490.

Lin, Peter Z, and Christopher M Meissner. 2021. “Persistent Pandemics.” Economics & Human Biology 43:101044.

Lin, Peter Zhixian, and Christopher M Meissner. 2020. A note on long-run persistence of public health outcomes in pandemics. Technical report. National Bureau of Economic Research.

Lo, Ming-Cheng M, and Hsin-Yi Hsieh. 2020. “The “Societalization” of pandemic unpreparedness: lessons from Taiwan’s COVID response.” American journal of cultural sociology 8 (3): 384–404.

Lokshin, Michael, Vladimir Kolchin, and Martin Ravallion. 2020. Scarred but Wiser: World War 2’s COVID Legacy. Working Paper, Working Paper Series 28291. National Bureau of Economic Research. doi:10.3386/w28291. http://www.nber.org/papers/w28291.

Ma, Junling, Jonathan Dushoff, and David JD Earn. 2011. “Age-specific mortality risk from pandemic influenza.” Journal of theoretical biology 288:29–34.

Mazumder, Bhashkar, Douglas Almond, Kyung Park, Eileen M Crimmins, and Caleb E Finch. 2010. “Lingering prenatal effects of the 1918 influenza pandemic on cardiovascular disease.” Journal of developmental origins of health and disease 1 (1): 26.

Nepomuceno, Marília R., Enrique Acosta, Diego Alburez-Gutierrez, José Manuel Aburto, Alain Gagnon, and Cássio M. Turra. 2020. “Besides population age structure, health and other demographic factors can contribute to understanding the COVID-19 burden.” Proceedings of the National Academy of Sciences 117 (25): 13881–13883. doi:10.1073/pnas.2008760117. eprint: https://www.pnas.org/doi/pdf/10.1073/pnas.2008760117. https://www.pnas.org/doi/abs/10.1073/pnas.2008760117.

Noymer, Andrew, and Michel Garenne. 2000. “The 1918 Influenza Epidemic’s Effects on Sex Differentials in Mortality in the United States.” Population and Development Review 26 (3): 565–581. doi:https://doi.org/10.1111/j.1728-4457.2000.00565.x. eprint: https://onlinelibrary.wiley.com/doi/pdf/10.1111/j.1728-4457.2000.00565.x. https://onlinelibrary.wiley.com/doi/abs/10.1111/j.1728-4457.2000.00565.x.

Papageorge, Nicholas W., Matthew V. Zahn, Michele Belot, Eline van den Broek-Altenburg, Syngjoo Choi, Julian C. Jamison, and Egon Tripodi. 2021. “Socio-demographic factors associated with self-protecting behavior during the Covid-19 pandemic.” Journal of Population Economics 34 (2): 691–738. ISSN: 1432-1475. doi:10.1007/s00148-020-00818-x. https://doi.org/10.1007/s00148-020-00818-x.

Piraino, Frank F, Edwin M Brown, and Edward R Krumbiegel. 1970. “Outbreak of Hong Kong influenza in Milwaukee, winter of 1968-69.” Public Health Reports 85 (2): 140.

Reichert, Tom, Gerardo Chowell, and Jonathan A McCullers. 2012. “The age distribution of mortality due to influenza: pandemic and peri-pandemic.” BMC medicine 10 (1): 1–15.

Sabeti, Pardis C, Emily Walsh, Steve F Schaffner, Patrick Varilly, Ben Fry, Holli B Hutcheson, Mike Cullen, Tarjei S Mikkelsen, Jessica Roy, Nick Patterson, et al. 2005. “The case for selection at CCR5-Δ 32.” PLoS Biol 3 (11): e378.

Saunders-Hastings, Patrick R, and Daniel Krewski. 2016. “Reviewing the History of Pandemic Influenza: Understanding Patterns of Emergence and Transmission.” Publisher: MDPI, Pathogens (Basel, Switzerland) 5 (4): 66. ISSN: 2076-0817. doi:10.3390/pathogens5040066. https://pubmed.ncbi.nlm.nih.gov/27929449.

SEER Program, National Cancer Institute, NIH. 2020. US Population Data. Underlying data provided by National Center for Health Statistics. Accessed May 15, 2020 at https://seer.cancer.gov/popdata/download.html#19.

Sette, Alessandro, and Shane Crotty. 2020. “Pre-existing immunity to SARS-CoV-2: the knowns and unknowns.” Nature Reviews Immunology 20 (8): 457–458.

Simonsen, Lone, Matthew J. Clarke, Lawrence B. Schonberger, Nancy H. Arden, Nancy J. Cox, and Keiji Fukuda. 1998. “Pandemic versus Epidemic Influenza Mortality: A Pattern of Changing Age Distribution.” The Journal of Infectious Diseases 178 (1): 53–60. ISSN: 0022-1899. doi:10.1086/515616. eprint: https://academic.oup.com/jid/article-pdf/178/1/53/2722505/178-1-53.pdf. https://doi.org/10.1086/515616.

Tai, Don Bambino Geno, Aditya Shah, Chyke A Doubeni, Irene G Sia, and Mark L Wieland. 2020. “The disproportionate impact of COVID-19 on racial and ethnic minorities in the United States.” Clinical Infectious Diseases.

Takahashi, Takehiro, and Akiko Iwasaki. 2021. “Sex differences in immune responses.” Publisher: American Association for the Advancement of Science Section: Perspective, Science 371 (6527): 347–348. ISSN: 0036-8075, 1095-9203, accessed January 29, 2021. doi:10.1126/science.abe7199. https://science.sciencemag.org/content/371/6527/347.

Taylor, Charles A, Christopher Boulos, and Douglas Almond. 2020. “Livestock plants and COVID-19 transmission.” Proceedings of the National Academy of Sciences 117 (50): 31706–31715.

U.S. Census Bureau. 2019a. All Sectors: County Business Patterns by Legal Form of Organization and Employment Size Class for U.S., States, and Selected Geographies: 2018. https://www2.census.gov/programs-surveys/cbp/data/2018/CB1800CBP.zip.

U.S. Census Bureau. 2019b. Commuting characteristics by sex, 2004-2018 American Community Survey 5-year estimates. https://data.census.gov/cedsci/table?tid=ACSST1Y2018.S0801.

U.S. Census Bureau. 2019c. Group quarters population by group quarters type, 2004-2018 American Community Survey 5-year estimates. https://data.census.gov/cedsci/table?q=group%5C%20quarters&tid=ACSST1Y2018.S2602.

U.S. Census Bureau. 2019d. Households and families, 2004-2018 American Community Survey 5-year estimates. https://data.census.gov/cedsci/table?tid=ACSST1Y2017.S1101.

U.S. Census Bureau. 2019e. Selected characteristics of health care coverage in the United States, 2004-2018 American Community Survey 5-year estimates. https://data.census.gov/cedsci/table?tid=ACSST1Y2018.S2701.

U.S. Department of Transportation. 2020. FAF4 Network Database and Flow Assignment: 2012 and 2045. https://ops.fhwa.dot.gov/freight/freight_analysis/faf/faf4/netwkdbflow/index.htm.

US Bureau of Economic Analysis. 2020. Table SAINC5: Personal Income by Major Component and Industry. Accessed May 15, 2020 at https://apps.bea.gov/iTable/iTable.cfm?reqid=70&step=1#reqid=70&step=1.

Wells, Chad R., Pratha Sah, Seyed M. Moghadas, Abhishek Pandey, Affan Shoukat, Yaning Wang, Zheng Wang, Lauren A. Meyers, Burton H. Singer, and Alison P. Galvani. 2020. “Impact of international travel and border control measures on the global spread of the novel 2019 coronavirus outbreak.” Proceedings of the National Academy of Sciences 117 (13): 7504–7509. doi:10.1073/pnas.2002616117. eprint: https://www.pnas.org/content/117/13/7504.full.pdf. https://www.pnas.org/content/117/13/7504.

Worby, Colin J, Sandra S Chaves, Jacco Wallinga, Marc Lipsitch, Lyn Finelli, and Edward Goldstein. 2015. “On the relative role of different age groups in influenza epidemics.” Epidemics 13:10–16.

Worobey, Michael, Guan-Zhu Han, and Andrew Rambaut. 2014. “Genesis and pathogenesis of the 1918 pandemic H1N1 influenza A virus.” Proceedings of the National Academy of Sciences 111 (22): 8107–8112.

Zeberg, Hugo, and Svante Pääbo. 2020. “The major genetic risk factor for severe COVID-19 is inherited from Neanderthals.” Nature 587 (7835): 610–612. ISSN: 1476-4687. doi:10.1038/s41586-020-2818-3. https://doi.org/10.1038/s41586-020-2818-3.

